# OpenClinicalAI: enabling AI to diagnose diseases in real-world clinical settings

**DOI:** 10.1101/2021.09.08.21263249

**Authors:** Yunyou Huang, Nana Wang, Suqin Tang, Li Ma, Tianshu Hao, Zihan Jiang, Fan Zhang, Guoxin Kang, Xiuxia Miao, Xianglong Guan, Ruchang Zhang, Zhifei Zhang, Jianfeng Zhan, for the Alzheimer’s Disease Neuroimaging Initiative

**Author notes:** Data used in preparation of this article were obtained from the Alzheimer’s Disease Neuroimaging Initiative (ADNI) database (http://adni.loni.usc.edu). As such, the investigators within the ADNI contributed to the design and implementation of ADNI and/or provided data but did not participate in analysis or writing of this report. A complete listing of ADNI investigators can be found at: http://adni.loni.usc.edu/wp-content/uploads/how_to_apply/ADNI_Acknowledgement_List.pdf.

## Abstract

This paper quantitatively reveals the state-of-the-art and state-of-the-practice AI systems only achieve acceptable performance on the stringent conditions that all categories of subjects are known, which we call closed clinical settings, but fail to work in real-world clinical settings. Compared to the diagnosis task in the closed setting, real-world clinical settings pose severe challenges, and we must treat them differently. We build a clinical AI benchmark named Clinical AIBench to set up real-world clinical settings to facilitate researches. We propose an open, dynamic machine learning framework and develop an AI system named OpenClinicalAI to diagnose diseases in real-world clinical settings. The first versions of Clinical AIBench and OpenClinicalAI target Alzheimer’s disease. In the real-world clinical setting, OpenClinicalAI significantly out-performs the state-of-the-art AI system. In addition, OpenClinicalAI develops personalized diagnosis strategies to avoid unnecessary testing and seamlessly collaborates with clinicians. It is promising to be embedded in the current medical systems to improve medical services.

**One-Sentence Summary:** We propose a clinical AI benchmark and an open, dynamic machine learning framework to enable AI diagnosis systems to land in real-world clinical settings.

## Introduction

Due to previous successive successes of AI in the clinical research field, AI is considered a promising technology to provide high-quality and low-cost diagnostic services (*1,2,3,4,5,6,7*). However, there is little evidence that these researches can be implemented into real-world clinical settings (in short, real-world settings) and improve medical services (*8, 9, 10*). Fig. 1, 2, 3 qualitatively and quantitatively reveal the state-of-the-art and state-of-the-practice AI systems only achieve acceptable performance on the stringent conditions. We call those stringent conditions closed clinical settings (in short, closed settings). The closed settings have the following primary assumptions: all categories of subjects are known a priori (*11*); the same diagnostic strategy is applied to all subjects, e.g., every subject requires a nuclear magnetic resonance scan (MRI) (*12*); the state-of-the-art AI systems can only be deployed at medical institutions that are able to execute the pre-prescribed diagnostic strategy (*4, 13, 14*). Vice versa, if the medical institution can not meet prerequisite conditions that are able to complete the pre-prescribed diagnostic strategy, the corresponding AI system can not be deployed. In this context, the diagnosis problem is a closed set recognition problem that is artificially simplified (*3, 4, 14, 5, 15*).

**Fig. 1:**
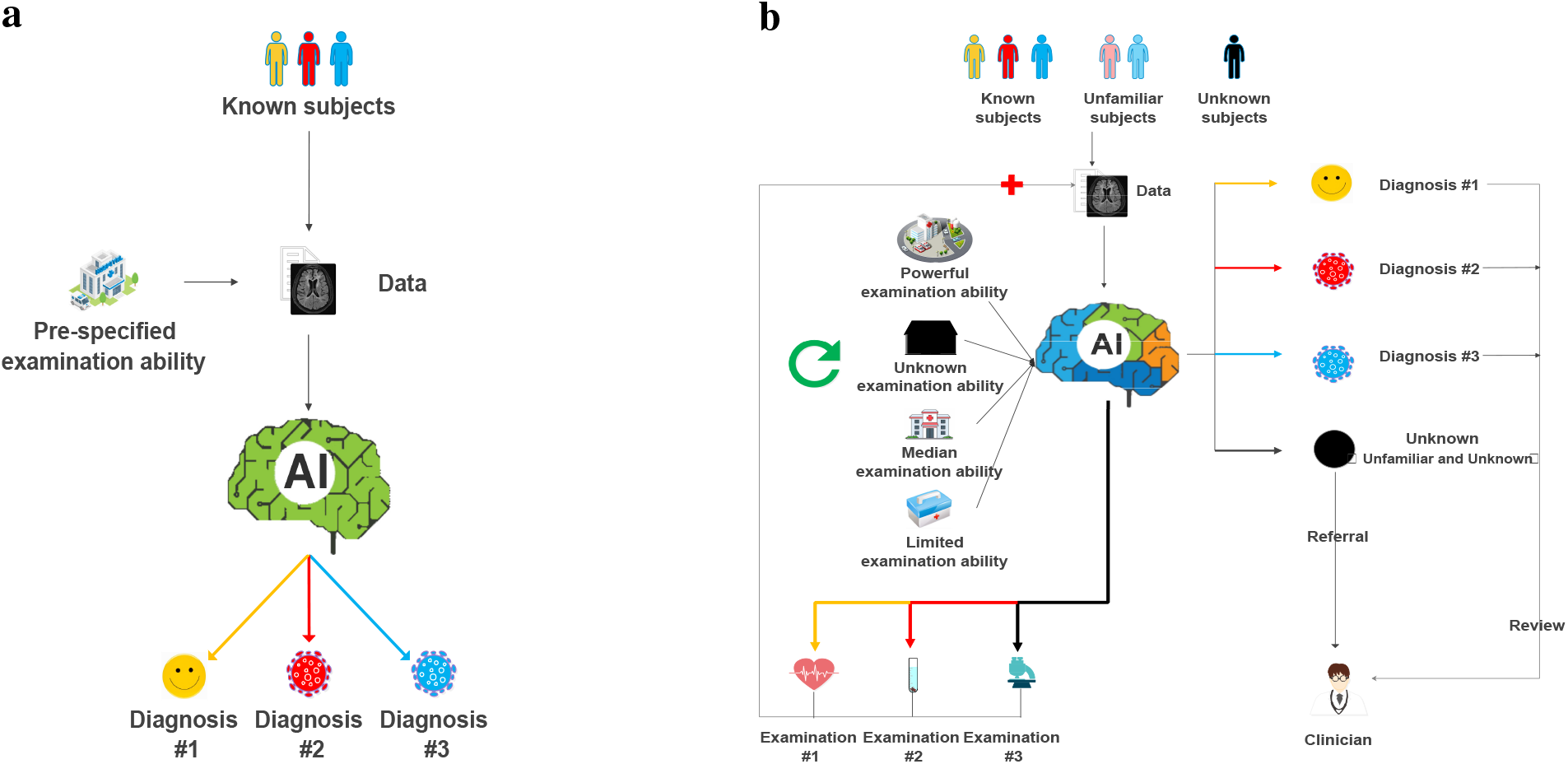
The workflow of the baseline clinical AI system and OpenClinicalAI. **a**, The workflow of the mainstream AI-based diagnostic systems for closed settings. The system only accepts subjects with pre-specified clinical states. First, the same pre-specified medical examinations will be executed by a medical institution with the pre-specified examination ability for every subject. And then, the system will calculate the probability of each pre-defined clinical state for the subject according to the examination. Finally, the system will take the clinical states with the maximum probability as the output and make the final diagnosis. **b**, The workflow of OpenClinicalAI. It can deal with different categories of subjects, including the unfamiliar and unknown categories of subjects during the development of the system. It can deploy in various medical institutions with different examination abilities from small-scale country clinics to large-scale hospitals. First, OpenClinicalAI will obtain the basic information of the subject and combine the history clinical information of the subject as input. Second, according to the input, OpenClinicalAI calculates the probability of each disease-related examination and each pre-defined clinical state, including the unknown clinical state. Third, for each pre-defined clinical state, if a clinical state’s possibility is greater than the specific threshold, then the clinical state is the final diagnosis of OpenClinicalAI, which will be sent to clinicians to review. Otherwise, go to the next step. Fourth, for each examination, if the probability of an examination is greater than the specified threshold and the medical institution can execute this examination, then obtain the examination data, add the data to the input of OpenClinicalAI, and go to step two. Otherwise, go to the next step. Fifth, for the medical institution with specific examination ability, select an executable routine examination with the least cost that has not been executed for the subject, add the examination data to the input of OpenClinicalAI, and go to step two. Otherwise, go to the next step. Finally, mark the subject without diagnosis as unknown and send them to clinicians to diagnose. Notably, the atypical subject with specified clinical states, unfamiliar and unknown subjects are marked as unknown and sent to the clinician for diagnosis.

**Fig. 2:**
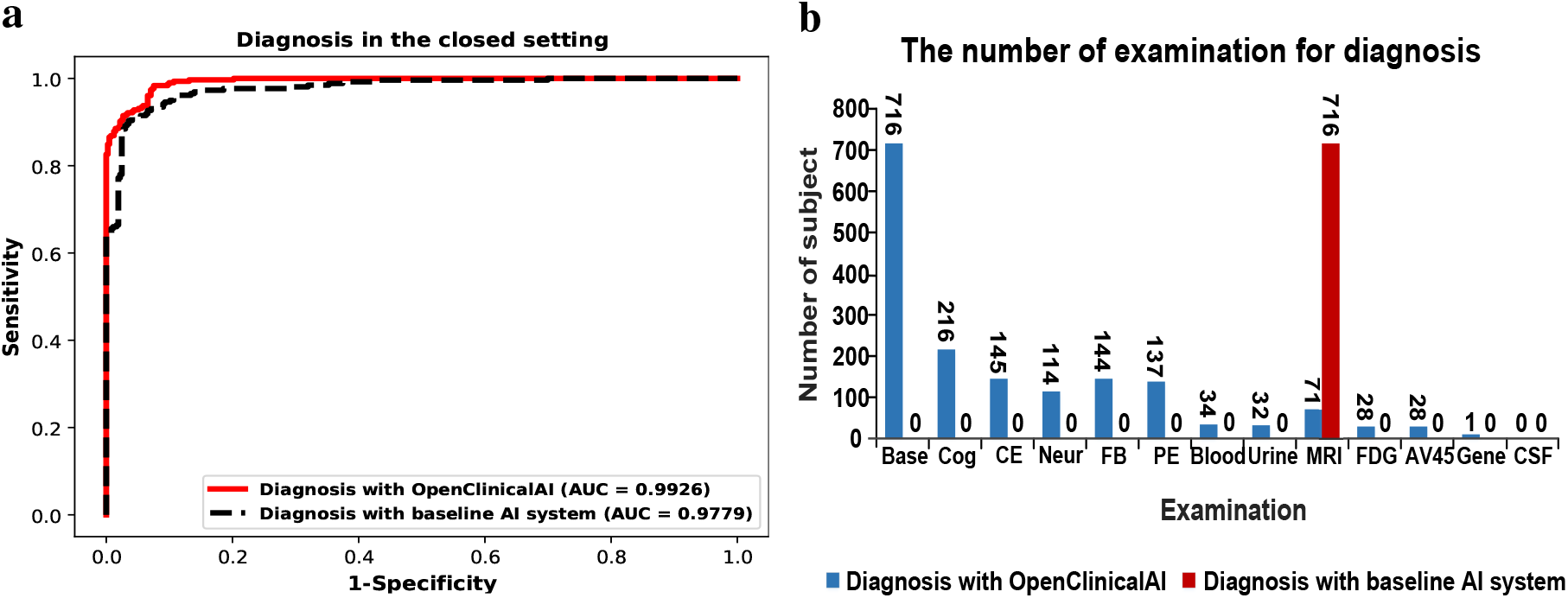
The performance of OpenClinicalAI with personalized strategies against the baseline system on Alzheimer’s disease diagnosis task in the closed setting. **a**, The ROC curves of two systems in the closed dataset. The red curve is the ROC curve of the baseline system, and it obtains an AUC score of 0.9779 (95% CI 0.9722-0.9827). The black curve is the ROC curve of OpenClinicalAI with various examination data, and it obtains an AUC score of 0.9926 (95% CI 0.9907-0.9945). **b**, The examination used during the AD diagnosis process. The baseline system consistently uses MRI data and historical data as the system input. In other words, every subject must have a nuclear magnetic resonance scan. OpenClinicalAI is able to develop and adjust the diagnosis strategies according to individual conditions and existing medical conditions during the diagnosis process, and only 71 subjects in the test set should have a nuclear magnetic resonance scan. Most subjects only need to have two or several simple examinations during the diagnosis process.

**Fig. 3:**
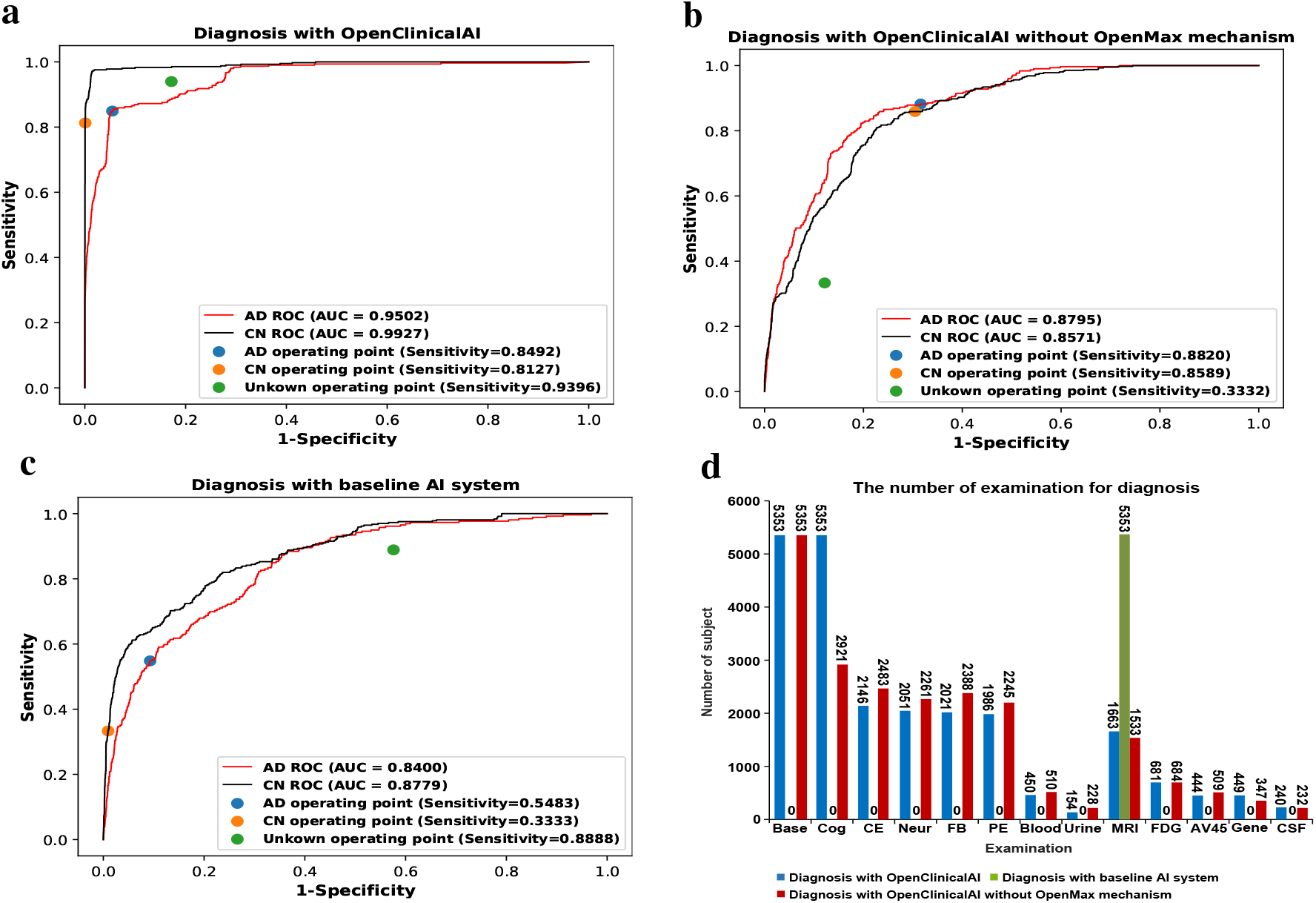
The performance of OpenClinicalAI with personalized strategies against the baseline system in the real-world setting. **a**, The ROC curves of OpenClinicalAI. It obtains two high AUC scores of 0.9502 (95% CI 0.9304-0.9662) and 0.9927 (95% CI 0.9854-0.9981) for AD and CN detection. The operating point of AI system is a group of score thresholds that separates positive and negative decisions of every category of the subject (0.95 for AD, 0.95 for CN, and 0.8 for unknown). On the operating point, OpenClinicalAI obtains the sensitivity of AD, CN, and unknown are 0.8492 (95% CI 0.7891-0.9051), 0.8127 (95% CI 0.7551-0.8667), and 0.9396 (95% CI 0.9290-0.9492) respectively. **b**, The ROC curves of OpenClinicalAI without an OpenMax mechanism. It obtains two AUC scores of 0.8795 (95% CI 0.8540-0.9038) and 0.8571 (95% CI 0.8331-0.8797) for AD and CN detection. On the operating point, the sensitivity of AD and CN are 0.8820 (95% CI 0.8282-0.9324) and 0.8589 (95% CI 0.8100-0.9056), respectively. However, the sensitivity of the unknown is only 0.3332 (95% CI 0.3133-0.3528). **c**, The ROC curves of the baseline system. The baseline system obtains two AUC scores of 0.8400 (95% CI 0.8055-0.8728) and 0.8779 (95% CI 0.8506-0.9025) for AD and CN detection. On the operating point, the unknown’s sensitivity is 0.8888 (95% CI 0.8753-0.9018). However, the sensitivity of AD and CN are only 0.5483 (95% CI 0.4604-0.6301) and 0.3333 (95% CI 0.2663-0.3979), respectively. **d**, The examination used during the AD diagnosis process. All subjects diagnosed by the baseline system require the nuclear magnetic resonance scan. The subject diagnosed by OpenClinicalAI without an OpenMax mechanism is similar to the subject diagnosed by OpenClinicalAI with an OpenMax mechanism. The selection of examination depends on the situation of the subject and existing medical conditions. Thus the examination number is not fixed.

Close settings are too ideal for real-world settings. The real-world setting is open with uncertainty and complexity. The subject in real-world settings is not all pre-known categories but contains many unknown and unfamiliar categories. Every subject is different, and there is no one-size-fits-all diagnosis strategy. Conditions of medical institutions are different and not pre-known, e.g., some hospitals have positron emission tomography (PET). In contrast, most of the other hospitals in underdeveloped areas are not equipped with PET. The diagnosis problem in real-world settings is an open set recognition problem (*16*).

Essentially, the diagnosis task in the closed setting is to find the optimal solution to classify different categories of subjects in a limited space (so-called supervised task) with the help of the ground truth of every subject. However, the real-world setting is open and puts the diagnosis task into unlimited space. Compared to the limited space of closed settings, the infinite space of real-world settings infinitely expands the scale of solving. Moreover, supervised learning will lose efficacy since some categories of subjects and their ground truth are unknown during the development of the AI model. Hence, the main problem of the diagnosis task in the real-world setting converts to efficiently locate the known subjects from the uncertain and complex real-world setting. Moreover, as shown in Fig. 2a and 3b,c, solving well the diagnosis task in the closed setting is not much help to solve the diagnosis task in the real-world setting. Compared to the diagnosis task in the closed setting, the diagnosis task in the real-world setting is a new and challenging task that we must treat differently.

This paper calls for turning medical AI attention from algorithmic research in closed settings to systematic study in real-world settings. Specifically, we construct a clinical AI benchmark named Clinical AIBench, which contains real-world and closed settings to promote the landing of AI in real-world settings. To tackle uncertainty and complexity in real-world settings, we propose an open, dynamic machine learning framework (Fig. S1) and a diagnostic system named OpenClinicalAI to embed in the current healthcare systems as shown in Fig. 1b.

The first versions of Clinical AIBench and OpenClinicalAI target Alzheimer’s disease (AD) as AD is an incurable disease that brings a heavy burden to our society (the total payment for individuals with AD or other dementias is estimated at 277 billion) (*17, 18, 19, 20*). Early and accurate AD diagnosis will result in the correct management of AD or other dementias, saving up to $7.9 trillion in medical and care costs (*19*). However, it is estimated that 28 million of the world’s 36 million people with dementia do not receive a diagnosis since the limited medical resources and experts, etc. (*21*).

The current version of Clinical AIBench includes two clinical settings, which are curated from a large enriched dataset Alzheimer’s disease neuroimaging initiative (ADNI): a closed setting and a real-world setting (*22*). OpenClinicalAI is composed of multiple independent parts, which can cooperate to handle unknown subjects in real-world settings, and dynamically adjust diagnosis strategies according to specific subjects and medical institutions. OpenClinicalAI provides an opportunity to embed the AI-based diagnostic system into the current healthcare systems to cooperate with clinicians to improve healthcare services.

In the real-world setting of Clinical AIBench, we evaluate the performance of OpenClinicalAI against the state-of-the-art AI diagnosis system. Our evaluations show that the performance of OpenClinicalAI exceeds that of the state-of-the-art AI diagnosis system in the real-world setting. Additionally, OpenClinicalAI can develop personalized diagnosis strategies for every subject in the real-world setting, maximizing the patient benefit.

## Results

### Clinical AIBench

Clinical AIBench contains real-world and closed settings to develop and evaluate the AI system designed for real-world settings. The first version targets Alzheimer’s disease. In this section, we focus on real-world settings.

The diagnosis in a real-world setting requires clinicians to use both individual clinical expertise and the best available external evidence, which is usually obtained by clinical examination, to make a clinical decision for every specific subject (*23*). It means that at least two main factors must be considered in the diagnosis task in real-world settings: the subject and the available clinical examination in the medical institution.

As shown in Fig. S2, the real-world setting is open with uncertainty and complexity. The primary characteristics of the real-world setting are as follows:

1. Real-world settings are open, and clinicians or AI systems often refer to unknown and unfamiliar categories. Thus, the subject’s categories are not all pre-known and familiar. A clinician has different expertise and may be unfamiliar with some diseases. In the real-world setting of Clinical AIBench, an unknown subject category means that it is not familiar to the clinician or AI system. Thus, we mark both unknown categories and unfamiliar categories as unknown. In this work, Clinical AIBench divides all mild cognitive impairment (MCI) and significant memory concern (SMC) subjects into the test set, which are unknown categories during the development of the AI system.
2. Subjects in real-world settings are under different situations. In this work, subjects with varying conditions are from 67 sites in two countries (Table S1). For every subject, data of all visits are included in Clinical AIBench (Table S2). The interval between two contiguous visits of a subject is usually more than six months.
3. Medical institutions in real-world settings have wildly different executive abilities of the examination. Not all the specific medical institutions and their specific executive abilities of the examination are pre-known. In this work, missing data for subjects are not be filled in the real-world setting of Clinical AIBench. In the real world, most of the subjects do not have all examination data categories. The purpose of the lack of specific category examination data is to keep the varied executive ability of the examination in different medical institutions. That is to say, in the real-world setting of Clinical AIBench, the lack of specific category examination data indicates that a medical institute lacks that examination ability.

Specifically, in this work, the examination data in ADNI is divided into 13 categories: base information (Base), cognition information (Cog), cognition testing (CE), neuropsychiatric information (Neur), function and behavior information (FB), physical neurological examination (PE), blood testing (Blood), urine testing (Urine), nuclear magnetic resonance scan (MRI), positron emission computed tomography scan with 18-FDG (FDG), positron emission computed tomography scan with AV45 (AV45), gene analysis (Gene), and cerebral spinal fluid analysis (CSF).

Details of the dataset in the real-world setting are as follows.

1. All subjects with labels in ADNI are included.
2. 85% AD and cognitively normal (CN) subjects are divided as the training set. 5% of AD and CN subjects are divided as the validation set. 20% AD and CN subjects, 100% MCI subjects, and 100% SMC are divided as the test set.
3. For every subject, different diagnosis strategies are combined according to the presence of different examination data, and the data of each diagnosis strategy forms a sample.

The test set is not accessible during the training of the AI system. In addition, since each subject may have multiple visits (each visit of the subject is treated as an independent subject), we stipulate that each subject’s visit data can only appear in one of the training set, validation set, and test set.

Sine previous AD diagnosis researches are developed in closed settings, the closed setting in Clinical AIBench is similar to the previous research (*24, 25, 26, 27, 28, 12, 29, 30, 31*). Only AD and CN subjects are included in the closed setting, and only the nuclear magnetic resonance instrument and historical medical records are available. 80% of subjects are divided as the training set, 5% of subjects are divided as the validation set, and 15% of subjects are divided as the test set.

### The performance of OpenClinicalAI on Alzheimer’s disease diagnosis

Ebrahimighahnavieh et al. and Tanveer et al. review many important works of Alzheimer’s disease diagnosis (*27, 28*). Most of these works are based on MRI data and transfer learning obtain the most excellent results. In addition, among the recent AI diagnosis researches, the transfer learning framework of the pre-trained model followed by a classifier achieves the state-of-the-art performance in many diagnosis tasks based on medical images (*14,1,32,33,3*). Thus, based on the state-of-the-art transfer learning framework and MRI data, we utilize a trained model named DenseNet201 (*34*) and a classifier called XGBoot (*35*) to develop an Alzheimer’s disease diagnosis AI system which we consider as the baseline system to compare against OpenClinicalAI in the rest of this paper.

We validate the effectiveness of OpenClinicalAI in two ways. First, we compare OpenClinicalAI to the baseline system in the closed setting. Second, we compare OpenClinicalAI to the baseline system in the real-world setting. Our comparison metrics are the area under the receiver operating characteristic (ROC) curve (AUC) and sensitivity. The larger the value of AUC and sensitivity are, the better the AI system is.

### The performance of OpenClinicalAI against the baseline system in the closed setting

To the best of our knowledge, all state-of-the-art and state-of-the-practice Alzheimer’s disease diagnosis AI researches are developed and evaluated in closed settings (*27, 28, 12, 29, 30, 31*). We firstly assess the baseline AI system in the closed setting, and then evaluate OpenClinicalAI in the same closed setting without the limitation of that only the nuclear magnetic resonance instrument and historical medical records are available.

As shown in Fig. 2 a, the baseline system obtains a high AUC score of 0.9779 (95% confidence interval (CI) 0.9722-0.9827), and there is not much room for promotion. OpenClinicalAI achieves an AUC score of 0.9926 (95% CI 0.9907-0.9945) and obtains the state-of-the-art performance. However, the essential improvement from the baseline system to OpenClinicalAI is that the latter can dynamically develop personalized diagnosis strategies according to specific subjects and medical institutions. As shown in Fig. 2 b, less than 10% of the subjects require a nuclear magnetic resonance scan, and most of the subjects only require harmless examination such as cognitive examination. We conclude OpenClinicalAI can avoid unnecessary examination for subjects and suit medical institutions with different examination abilities ^1^.

### The performance of OpenClincalAI against the baseline system in the real-world setting

Our goal is to develop an AI diagnosis system that can be embedded in the current medical system and cooperated with clinicians. In this work, if the predicted probability of the AD or CN is smaller than the probability threshold (0.95), the subject will be marked as unknown and referral to the clinician. For comparison, we use the same baseline system discussed above. In addition, we also consider OpenClinicalAI without an OpenMax mechanism (Algorithm S2,3) as the comparison system (*11*).

As shown in Fig. 3a, b, and c, compared to the baseline system, OpenClinicalAI demonstrates a significant improvement in the AUC of identification of AD subjects (+0.1102) and the AUC of identification of CN subjects (+0.1148). It is worth noting that OpenClinicalAI has a vast improvement in the sensitivity of AD, CN, and unknown on the operating point.

For the baseline system, the sensitivity of known (AD and CN) subjects is low. The sensitivity of AD is just 0.5483 (95% CI 0.4604-0.6301), and the sensitivity of CN is just 0.3333(95% CI 0.2663-0.3979). It indicates that most known subjects will be marked as unknown and sent to the clinician for diagnosis. Moreover, the sensitivity of unknown subjects is 0.8888(95% CI 0.8753-0.9018), meaning 11.12% of unknown subjects will be misdiagnosed. In addition, the baseline system requires that every subject has a nuclear magnetic resonance scan, and every medical institution that deploys the baseline system must be equipped with a nuclear magnetic resonance apparatus.

For OpenClinicalAI without an OpenMax mechanism, the sensitivity of known (AD and CN) subjects is as good as OpenClinicalAI with an OpenMax mechanism. In contrast, the sensitivity of unknown subjects is much worse than OpenClinicalAI with an OpenMax mechanism. It means most unknown subjects will be misdiagnosed, and it is unendurable in real-world settings.

OpenClinicalAI diagnoses most of the known (AD and CN) subjects correctly, marks most of the rest as unknown, and sends them to the clinician for further diagnosis. Besides, most unknown subjects are correctly identified, and the misdiagnosis of unknown subjects is only 6.04%. It means that OpenClinicalAI has enormous potential application value to implement in real-world settings. In addition, as shown in Fig. 3d, similar to the behaviors of OpenClinicalAI in the closed setting, OpenClinicalAI can develop and adjust diagnosis strategies for every subject dynamically in the real-world setting. Only a small part of subjects require a nuclear magnetic resonance scan and more costs (economy and harm) examinations.

### Development of diagnosis strategies

For every subject, firstly, OpenClinicalAI will acquire the base information of the subject. Secondly, OpenClinicalAI will give a final diagnosis or receive other examination information according to the current data of the subject. Thirdly, repeat the previous step until the diagnosis is finalized or there is no further examination.

As shown in Fig. 4a, diagnosis strategies of subjects are not the same (Table S3). OpenClinicalAI dynamically develops 35 diagnosis strategies according to different subject situations and all 40 examination abilities in the test set (Table S4). For the known (AD and CN) subjects, as shown in Fig. 4b, and c, most of the subjects require low-cost examinations (such as cognition examination (CE)). A small part of subjects requires high-cost examinations (such as cerebral spinal fluid analysis (CSF)). For unknown subjects, as shown in Fig. 4d, different from the diagnosis of known (AD and CN) subjects, identifying unknown subjects is more complex and more dependent on high-cost examinations. The reason for the above phenomenon is that according to the mechanism of OpenClinicalAI, it will do its best to distinguish whether the subject belongs to the known categories. When it fails, OpenClinicalAI will mark the subject as unknown. It means that the unknown subject will undergo more examinations than the known subject. The details of the high-cost examinations requirement are as follows.

**Fig. 4:**
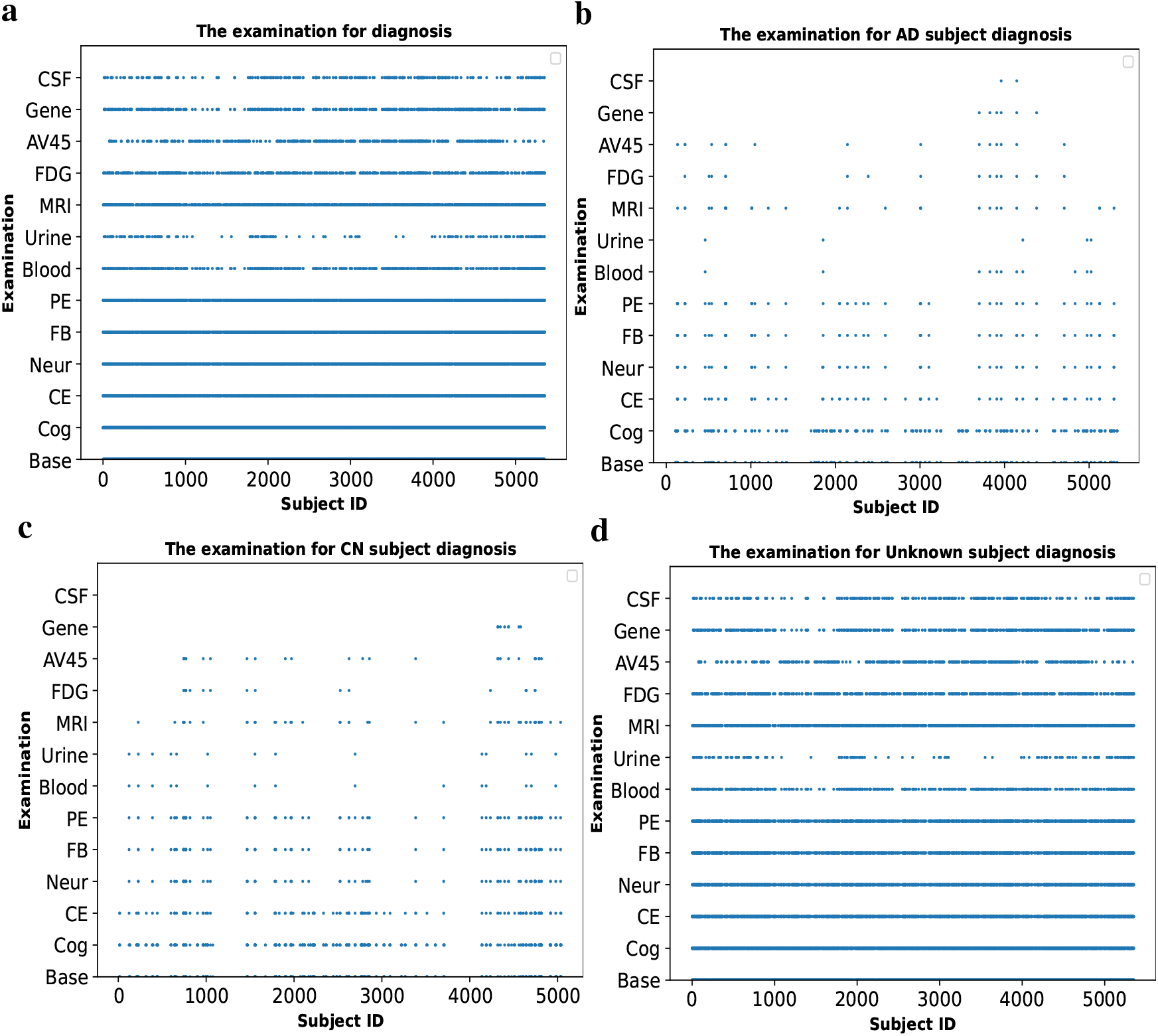
Diagnosis strategies for subjects. **a**, Diagnosis strategies for all subjects. Due to OpenClinicalAI developing and adjusting the examination for each subject, the selection of examinations for subjects is not the same. **b**, Diagnosis strategies for AD subjects. Compared to the high-cost examination, OpenClinicalAI pays more attention to the subject’s basic information, cognitive, mental, behavioral, and physical examination information for the AD subject. In contrast, biochemical testing, imaging, and genetic data are less considered. **c**, Diagnosis strategies for CN subjects. The behaviors of OpenClinicalAI for CN recognition are similar to those for AD diagnosis, and the difference between those behaviors is that more examinations are required to identify the CN subject. **d**, Diagnosis strategies for unknown (MCI and SMC) subjects. Compared to the known subject recognition, identifying unknown subjects is more complicated, and more examinations are required.

1. 33.94% of unknown subjects require a nuclear magnetic resonance scan (that of the known subject is 12.43%).
2. 13.95% of unknown subjects require a positron emission computed tomography scan with 18-FDG (that of the known subject is 4.75%).
3. 8.67% of unknown subjects require a positron emission computed tomography scan with AV45 (that of the known subject is 5.87%).
4. 9.38% of unknown subjects require a gene analysis (that of the known subject is 1.96%).
5. 5.13% of unknown subjects require a cerebral spinal fluid analysis (that of the known subject is 0.28%).

### Potential clinical applications

OpenClinicalAI enables that the AD diagnosis system can be implemented in uncertain and complex clinical settings to reduce the workload of AD diagnosis and minimize the cost of subjects.

To identify the known (AD and CN) subject with high confidence, the operating point of OpenClinicalAI is running with a high decision threshold (0.95). For the test set, OpenClinicalAI achieved a accuracy value of 92.47% (95% CI 91.36%-93.44%), AD sensitivity value of 84.92% (95% CI 78.91%-90.51%), CN sensitivity value of 81.27% (95% CI 75.51%-86.67%) while retaining an unknown sensitivity value of 93.96% (95% CI 92.90%-94.92%). In addition, OpenClinicalAI can cooperate with the senior clinician to identify the known subject. In this work, 15.08% (95% CI 9.49%-21.09%) of AD subjects and 18.73% (95% CI 13.33%-24.49%) of CN subjects are marked as unknown and sent to senior clinicians to diagnose. The work pattern is significant for the undeveloped area, which is a promising way to connect developed areas and undeveloped areas to reduce the workload, improve the overall medical services, and promote medical equity. To minimize the subject cost and maximize the subject benefit, OpenClinicalAI dynamically develops personalized diagnosis strategies for the subject according to the subject’s situation and existing medical conditions.

For the subject, OpenClinicalAI will judge whether it can finalize the subject’s diagnosis according to the currently obtained information of subjects. If the current data of the subject is not enough to support OpenClinicalAI to make a diagnosis, it will recommend the most suitable further examination for the subject. It will mitigate the over-testing plight, minimize the subject cost, and maximize the subject benefits. For the test set, 35 different diagnosis strategies are applied to the subject by OpenClinicalAI (Table S3). The details of the high-cost examination are as follows.

1. 31.07% of subjects require a nuclear magnetic resonance scan.
2. 12.72% of subjects require a positron emission computed tomography scan with 18-FDG.
3. 8.29% of subjects require a positron emission computed tomography scan with AV45.
4. 8.39% of subjects require a gene analysis.
5. 4.48% of subjects require a cerebral spinal fluid analysis.

For the medical institution, before the system recommends an examination for a subject, OpenClinicalAI will inquire whether the medical institution can execute the examination. Suppose the medical institution cannot perform the examination. In that case, OpenClinicalAI will recommend other examinations until the current information of the subject is enough to support it to make a diagnosis or until all common examinations have been suggested and the subject is marked as unknown. It enables that OpenClinicalAI is able to deploy in the different medical institutions with various examination abilities. In this work, OpenClinicalAI diagnoses subjects on 40 conditions of medical institutions (Table S4). In addition, for the subject of the test set, due to lack of the information of recommended examinations (which may be equal to the medical institution not having the ability to execute the recommended examination), OpenClinicalAI adjusts the diagnostic strategies 14654 times.

## Discussion

Currently, the media overhype the AI assistance diagnosis system. However, it is far from being mature to be implemented in real-world clinical settings. Many clinicians are gradually losing faith in the medicine AI (*36, 37, 9, 38, 39, 40*). Similar to the first trough of AI, the high expectation and unsatisfactory practical implementation of medical AI may severely hinder the development of medical AI. In addition, compared performances of state-of-the-art AI systems on stringent conditions and real-world settings, solving well the diagnosis task on stringent conditions is not much help to solve the diagnosis task in the real-world setting. It is time to draw the attention from the pure algorithm research in closed settings to systematic study in real-world settings, focusing on the challenge of tackling the uncertainty and complexity of real-world settings. In this work, we propose an open, dynamic machine learning framework to make the AI diagnosis system can directly deal with the uncertainty and complexity in the real-world setting. Based on our framework, an AD diagnostic system demonstrates huge potentiality to implement in the real-world setting with different medical environments to reduce the workload of AD diagnosis and minimize the cost of the subject.

Although many AI diagnostic systems have been proposed, how to embed these systems into the current health care system to improve the medical service remains an open issue (*2, 41, 42, 43*). OpenClinicalAI provides a reasonable way to embed the AI system into the current health care system. OpenClinicalAI can collaborate with clinicians to improve the clinical service quality, especially the clinical service quality of undeveloped areas. On the one hand, OpenClinicalAI can directly deal with the diagnosis task in the uncertain and complex real-world setting. On the other hand, OpenClinicalAI can diagnose typical patients of known subjects, while sending those challenging or atypical patients of known subjects to the clinicians for diagnosis. Although AI technology is different from traditional statistics, the model of the AI system still learns patterns from training data. For typical patients, the model is easy to understand patterns from patients, while it is challenging to learn patterns for atypical patients. Thus, every atypical and unknown patient is needed to treat by clinicians especially. In this work, most of the known subjects are diagnosed by OpenClinicalAI, and the rest are marked as unknown and sent to the senior clinician.

Over-testing has always been a concern and has been exacerbated in current AI-based diagnostic systems (*44, 45*). As samples, the systems proposed by Lu et al., Ding et al., and Liu et al. achieved state-of-the-art performance. At the same time, they required every subject to have a positron emission computed tomography scan, which is unnecessary for most subjects in real-work settings (*46, 31, 47*). However, OpenClinicalAI enables AI systems able to develop personalized diagnosis strategies to avoid unnecessary testing. OpenClinicalAI provides a possible way that can effectively reduce over-testing under strict supervision.

Notably, the experiment of this work does not contain a comparison with clinicians. There are two main reasons. First, OpenClinicalAI obtains an AUC value of 0.9927 (95% CI 0.9854-0.9981) in the closed setting. It is very close to the ground truth and unnecessary compared to clinicians. Second, the diagnosis patterns in real-world settings aim to diagnose typical patients of known subjects (which is usually easier to diagnose) and send atypical patients of known subjects (which are generally difficult to diagnose) and unknown subjects to clinicians. The task of OpenClinicalAI is quite different from that one of clinicians. Unlike current AI-based diagnostic systems, OpenClinicalAI performs as a new part of the whole healthcare system instead of replacing the role of clinicians. Therefore, it is not necessary to compare OpenClinicalAI to clinicians.

Although OpenClinicalAI is promising to impact the future research of the diagnosis system, several limitations remain. First, the prospective clinical studies of diagnosis of Alzheimer’s disease will be required to prove the effectiveness of our system. Second, the data of collection and processing are required to follow the standards of ADNI.

## Data Availability

The data from Alzheimer's Disease Neuroimaging Initiative was used under license for the current study. Applications for access to the dataset can be made at http://adni.loni.usc.edu/data-samples/access-data/. All original code has been deposited at the website https://www.benchcouncil.org/ and is publicly available as of the date of publication.

## Acknowledgments

We thank Weibo Pan and Fang Li for downloading the raw data sets from Alzheimer’s Disease Neuroimaging Initiative.

## Author contributions

Y.H. conceptualized the study, designed the models, wrote the codes, collected and analyzed the data, and wrote the manuscript. N.W., S.T., L.M, T.H., and Z.J. conceptualized the study and revised the manuscript. F.Z., G.K., X.M, X.G, and R,Z. collected and analyzed the data. Z.Z., and J.Z. directed the project and revised the manuscript.

## Data and materials availability

The data from Alzheimer’s Disease Neuroimaging Initiative was used under license for the current study. Applications for access to the dataset can be made at http://adni.loni.usc.edu/data-samples/access-data/. All original code has been deposited at the website BenchCouncil and is publicly available as of the date of publication.

## Supplementary materials

Materials and Methods

Figs. S1 to S3

Tables S1 to S6

Algorithms S1 to S4

References *(48-70)*

## Footnotes

1 Different hospitals have various clinical settings, such as community hospitals without nuclear magnetic resonance machines, big hospitals with multiple facilities.

## References

1. A. Esteva, et al., Nature 542, 115 (2017).

2. S. M. McKinney, et al., Nature 577, 89 (2020).

3. D. S. Kermany, et al., Cell 172, 1122 (2018).

4. J. De Fauw, et al., Nature medicine 24, 1342 (2018).

5. K. Ning, et al., Neurobiology of aging 68, 151 (2018).

6. Z. Tang, et al., Nature communications 10, 1 (2019).

7. C. Lian, M. Liu, Y. Pan, D. Shen, IEEE Transactions on Cybernetics (2020).

8. J. He, et al., Nature medicine 25, 30 (2019).

9. P. Brocklehurst, et al., The Lancet 389, 1719 (2017).

10. M. Roberts, et al., Nature Machine Intelligence 3, 199 (2021).

11. A. Bendale, T. Boult, Proceedings of the IEEE conference on computer vision and pattern recognition (2015), pp. 1893–1902.

12. S. Qiu, et al., Brain 143, 1920 (2020).

13. J. J. Titano, et al., Nature medicine 24, 1337 (2018).

14. H. Lee, et al., Nature biomedical engineering 3, 173 (2019).

15. X. Mei, et al., Nature medicine 26, 1224 (2020).

16. C. Geng, S.-j. Huang, S. Chen, IEEE transactions on pattern analysis and machine intelligence (2020).

17. L. E. Hebert, L. A. Beckett, P. A. Scherr, D. A. Evans, Alzheimer Disease & Associated Disorders 15, 169 (2001).

18. L. E. Hebert, J. Weuve, P. A. Scherr, D. A. Evans, Neurology 80, 1778 (2013).

19. A. Association, et al., Alzheimer’s & Dementia 14, 367 (2018).

20. C. S. Frigerio, et al., Cell reports 27, 1293 (2019).

21. M. Prince, R. Bryce, C. Ferri (2018).

22. S. G. Mueller, et al., Neuroimaging Clinics 15, 869 (2005).

23. D. L. Sackett, W. M. Rosenberg, J. M. Gray, R. B. Haynes, W. S. Richardson, BMJ 312, 71 (1996).

24. H. Li, et al., Alzheimer’s & Dementia (2019).

25. H. Choi, et al., EBioMedicine 43, 447 (2019).

26. T. Zhou, M. Liu, K.-H. Thung, D. Shen, IEEE transactions on medical imaging (2019).

27. M. A. Ebrahimighahnavieh, S. Luo, R. Chiong, Computer methods and programs in biomedicine 187, 105242 (2020).

28. M. Tanveer, et al., ACM Transactions on Multimedia Computing, Communications, and Applications (TOMM) 16, 1 (2020).

29. R. Sharma, T. Goel, M. Tanveer, S. Dwivedi, R. Murugan, Applied Soft Computing 106, 107371 (2021).

30. M. Tanveer, et al., IEEE Journal of Biomedical and Health Informatics (2021).

31. Y. Ding, et al., Radiology 290, 456 (2019).

32. P. Tschandl, et al., Nature Medicine 26, 1229 (2020).

33. R. Poplin, et al., Nature Biomedical Engineering 2, 158 (2018).

34. G. Huang, Z. Liu, L. Van Der Maaten, K. Q. Weinberger, Proceedings of the IEEE conference on computer vision and pattern recognition (2017), pp. 4700–4708.

35. T. Chen, C. Guestrin, Proceedings of the 22nd acm sigkdd international conference on knowledge discovery and data mining (2016), pp. 785–794.

36. M. van Assen, L. J. Cornelissen, Jacc-Cardiovascular imaging 13, 1172 (2020).

37. C. G. Weaver, F. A. McAlister, Canadian Journal of Cardiology (2021).

38. J. H. Chen, S. M. Asch, The New England journal of medicine 376, 2507 (2017).

39. T. M. Maddox, J. S. Rumsfeld, P. R. Payne, Jama 321, 31 (2019).

40. H. T. Head, Bmj p. 363 (2018).

41. C.-Y. Kuo, H.-M. Chiu, Journal of Gastroenterology and Hepatology 36, 267 (2021).

42. J. Schneider, M. Agus, arXiv preprint 2103.01149 (2021).

43. J. Bullock, A. Luccioni, K. H. Pham, C. S. N. Lam, M. Luengo-Oroz, Journal of Artificial Intelligence Research 69, 807 (2020).

44. M. OKeeffe, et al., JAMA Internal Medicine 181, 865 (2021).

45. J. W. OSullivan, et al., BMJ open 8, e018557 (2018).

46. D. Lu, et al., Medical image analysis 46, 26 (2018).

47. M. Liu, D. Cheng, W. Yan, A. D. N. Initiative, et al., Frontiers in neuroinformatics 12, 35 (2018).

