## Supplementary materials for "OpenClinicalAI: enabling AI to diagnose diseases in real-world clinical settings"

#### This PDF file includes:

Materials and Methods

Figs. S1 to S3

Tables S1 to S6

Algorithms S1 to S4

References *(48-70)*

#### Materials and Methods

##### Human subjects

Data used in the preparation of this article were obtained from the Alzheimer's Disease Neuroimaging Initiative (ADNI) database (`http://adni.loni.usc.edu`). The ADNI was launched in 2003 as a public-private partnership, led by Principal Investigator Michael W. Weiner,MD. For up-to-date information, see `http://www.adni-info.org`.

The data is collected from 67 sites in the United States and Canada (*48, 49, 50, 51*). The subject in the dataset aged between $54.4$ and $91.4$ at the first visit. The interval of the subject follow-up is usually greater than $6$ months. Generally, the longer the follow-up time is, the longer the interval is. The first visit is marked as bl, and the other visit is marked as m$\underline{xx}$ according to the time (For example, the visit takes place six months after the first visit is marked as m06). Detailed characteristics of the subject are shown in Table S1,2.

#### Dataset

The data contains study data, image data, genetic data compiled by ADNI between 2005 and 2019. Considering the commonly used examinations and the concerned examinations in AD diagnosis by the clinician, 13 categories of data are selected.

(1) Base information, usually obtained through consultation, includes demographics, family history, medical history, symptoms.

(2) Cognition information, usually obtained through consultation and testing, includes Alzheimer's Disease Assessment Scale, Mini-Mental State Exam, Montreal Cognitive Assessment, Clinical Dementia Rating, Cognitive Change Index.

(3) Cognition testing, usually obtained through testing, includes ANART, Boston Naming Test, Category Fluency-Animals, Clock Drawing Test, Logical Memory-Immediate Recall, Logical Memory-Delayed Recall, Rey Auditory Verbal Learning Test, Trail Making Test.

(4) Neuropsychiatric information, usually obtained through consultation, includes Geriatric Depression Scale, Neuropsychiatric Inventory, Neuropsychiatric Inventory Questionnaire.

(5) Function and behavior information, usually obtained through consultation, includes Function Assessment Question, Everyday Cognitive Participant Self Report, Everyday Cognition Study Partner Report.

(6) Physical, neurological examination, usually obtained through testing, includes Physical Characteristics, Vitals, neurological examination.

The rest of the examinations include blood testing, urine testing, nuclear magnetic resonance scan, positron emission computed tomography scan with 18-FDG, positron emission computed tomography scan with AV45, gene analysis, and cerebral spinal fluid analysis. It is worth noting that not all categories of information are obtained for a subject's visit, and the information on each type is often incomplete.

All subjects with labels containing at least one of the above categories of information are considered in this study. Two thousand one hundred twenty-seven subjects with 9593 visits are included in our work. A subject in a visit may require different categories of examination. Every combination of those examinations represents a diagnosis strategy. Thus, for the subject, $443795$ strategies are generated. These AD and CN subjects are randomly assigned to the training, validation, and test set. The training set contains 1025 subjects with 3986 visits and generates 180682 strategies. In the training set, 587 subjects with 1781 visits are AD and develop 80022 strategies, 466 subjects with 2205 visits are CN, and generate 100660 strategies. The validation set contains 73 subjects with 254 visits and generates 11898 strategies. In the validation set, 44 subjects with 127 visits are AD and develop 6008 strategies, 31 subjects with 127 visits are CN, and generate 5890 strategies. The test set contains 1460 subjects with 5353 visits. In the test set, 109 subjects with 305 visits are AD, 92 subjects with 411 visits are CN, 1082 subjects with 4357 visits are MCI, 280 subjects with 280 visits are SMC. Notably, the label of a subject may be different in other visits.

#### Randomization and blinding.

AD and CN subjects as known categories of subjects are randomized into training, validation, and test sets by applying a random function provided by the Python3 tool. The assignment is determined by a float value generated by a random function. We assign subjects whose values are [0,0.8) into the training set, assign subjects whose values are [0.8,0.85) into the validation set, assign subjects whose values are [0.85,1] into the test set. The data of visits belong to the same subject are only allowed to appear in the same set. MCI and SMC subjects as unknown categories of subjects are directly into the test set. During the development of the AI system, the test set is inaccessible.

#### Data preparation.

For each category of study data, if it contains more than one sub-category of data, concatenate all of the sub-category data by RID (The ID of the subject) and VISCODE (The mark of the subject's visit). For the medical image, we first convert the data from the DICOM format to the NIfNI format by the dcm2nii library. Second, register the image by ant library (*52, 53, 54*). Third, convert the 3D image to 2D slices and convert the image from gay to RGB. Finally, a trained model named DenseNet201 is used to extract the features of the 2D slices (*34*). For the genetic data, we extract 70 single nucleotide polymorphisms (SNP), which are very relating to the AD ( Table S5), and use one-hot code to represent each SNP (*55,56,57*). This work proposes a unified data representation framework, since the different dimensions of each category of data, the number of data categories included in each visit is different, and the number of history visits included in each subject is also different. We present an examination category in the subject's visit by an array with a shape of $1 \times 2090$. The shape of our data is $n \times 2090$, $n$ is the number of categories of data for the subject ( Fig. S3).

#### The propose model

Our model consists of five parts: $Encoder\_1$, $Decoder$, $Classifier\_1$, $Encoder\_2$, and $Classifier\_2$ ( Fig. S1 ). We name the model consisting of $Encoder\_1$, $Decoder$, and $Classifier\_1$ as $sub\_model\_1$, which can identify the subject from open clinical settings (*16, 58, 11*). We name the model consisting of $Encoder\_2$, and $Classifier\_2$ as $sub\_model\_2$, which can dynamically develop and adjust the diagnosis strategy according to the situation of subjects and existing medical conditions.

#### Loss function

The $sub\_model\_1$ is a multi-task learning model, which simultaneously optimizes the model's disease diagnosis and data reconstruction ability. The data reconstruction task can improve the diagnosis ability of the model in the open world (*58*). The loss function of the model is $Loss = 0.65 * loss_{diagnosis} + 0.35 * loss_{reconstruction}$. The $loss_{diagnosis}$ is categorical cross-entropy, and the $loss_{reconstruction}$ is mean squared logarithmic error. The $sub\_model\_2$ is also a multi-task learning model, which simultaneously optimizes the 12 examinations whether should be selected as the next examination for the subject. We introduce a loss function that combines the BCE loss function and weighs losses with uncertainty (*59, 14*). The modified loss function is given by equations 1:

$$\begin{gathered} Loss = -\sum_{i=2}^{k} \frac{1}{2\delta_i^2}(\gamma_P^i y^i log\hat{y}^i + \gamma_N^i (1-y^i) log(1-\hat{y}^i)) + log\delta_i \\ \gamma_P^i = \frac{|P^i|+|N^i|}{2|P^i|} \qquad \gamma_N^i = \frac{|P^i|+|N^i|}{2|N^i|} \end{gathered} \tag{1}$$

where $|P^i|$ is the total number of $ith$ examinations as the subsequent examination, $|N^i|$ is the total number of other examinations as the following examination. $\delta_i$ is an observation noise scalar of the output of $ith$ examination (*59*).

#### Label examination

Although researchers have made many efforts on the interpretability and internal logic of deep learning, the current behavior of deep learning is still tricky to understand (*60, 61*). We do not know whether the diagnosis strategy of the AI model needs to be consistent with human experts. Thus, it is unnecessary to label the subsequent examination of the current examination strategy by the clinician and train a model to simulate the clinician's behavior. In this work, the following examination label is labeled by the examination label algorithm ( Algorithm S1 ). The subsequent examination for the subject is determined by whether this examination makes the prediction model ($sub\_model\_1$) obtain a greater predicted probability for the correct category and smaller predicted probabilities for other categories.

#### OpenMax

OpenMax is a modified SoftMax layer that adopted the concept of Meta-Recognition (*62, 11, 63*). OpenMax uses the distance between the activation vector (AV) of the sample and the mean activation vector (the mean computed over only the correctly classified training examples) to identify the unknown categories of the subject (*11*). The deep learning network can be regarded as a feature extractor, and the output of the AV layer can be regarded as characteristics of the sample. However, the AV layer usually only retains the most relevant features to the classification task, and the features related to the unknown category are not guaranteed to be retained. To alleviate this problem, we replaced the output of the AV layer with the abnormal patterns of 14 selected indicators of known categories according to the Alzheimer's Diagnosis guidelines to improve the performance of the AI model (*64,65,66,67*) ( Table S6 ). The modified OpenMax by abnormal patterns is shown in Algorithm S2,3.

#### Model training.

The training of our model consists of two stages. The first stage is training the $sub\_model\_1$, in which the $Classifier\_1$ uses SoftMax layer as the output layer. The dimension of the output of the $sub\_model\_1$ in this training stage is 2, corresponding to AD and CN. After training the $sub\_model\_1$, a modified OpenMax layer, which estimates the probability of an input being an unknown class, is used to replace the SoftMax layer (*11*). The dimension of the output of $sub\_model\_1$ in the prediction stage is 3, corresponding to AD, CN, and unknown. According to prediction probabilities of subjects by the $sub\_model\_1$, every examination strategy in the training set and validation set is labeled by the Algorithm S1. The second stage is training the $sub\_model\_2$, the input of the $sub\_model\_2$ contains raw data and the prediction probability, the dimension of the output of the $sub\_model\_2$ is 12, which respectively correspond to 12 categories of examination. The model was optimized using mini-batch stochastic gradient descent with Adam and a base learning rate of 0.0005 (*68*). The experiments are conducted on a Linux server equipped with Tesla P40 and Tesla P100 GPU.

Due to the historical information has a significant influence on the diagnosis of Alzheimer's disease, there is a vast difference between the diagnosis of Alzheimer's disease at first visit without historical information and other visits with historical data. Therefore, based on the above model training method, we additionally trained a model for diagnosing Alzheimer's disease at the first visit based on the subject's data at the first visit.

#### Prediction

Unlike the other state-of-the-art AI models, predictions of our model are dynamic. The prediction algorithm comprehensively considers the situation of the subject, the condition of the medical institution, and the ability of our model to dynamically adjust the diagnosis strategy ( Algorithm S4). Firstly, our model will generate the probability for every category (AD vs.

CN vs. Unknown) according to the current input data of the subject. Second, if the probability of categories exceeds thresholds (AD $0.95$, CN $0.95$, unknown $0.8$), output the corresponding label. Otherwise, adjust the examination strategy by selecting the subsequent examination according to the situation of the subject and the medical institution, and go to the first step. Finally, if all diagnostic strategies are tried, the model still cannot obtain the probability of exceeding the threshold and then outputs unknown.

#### Statistical analysis

To evaluate the evaluation index of the AI model, a non-parametric bootstrap method is applied to calculate the confidence intervals (CI) for the evaluation index (*69*). In this work, we calculate $95\%$ CI for every evaluation index. We randomly sample $2500$ cases from the test set and evaluated the AI model by the sampled set for every evaluation index. $2000$ repeated trials are executed, and $2000$ values of the evaluation index are generated. The 95% CI is obtained by the 2.5 and 97.5 percentiles of the distribution of the evaluation index values.

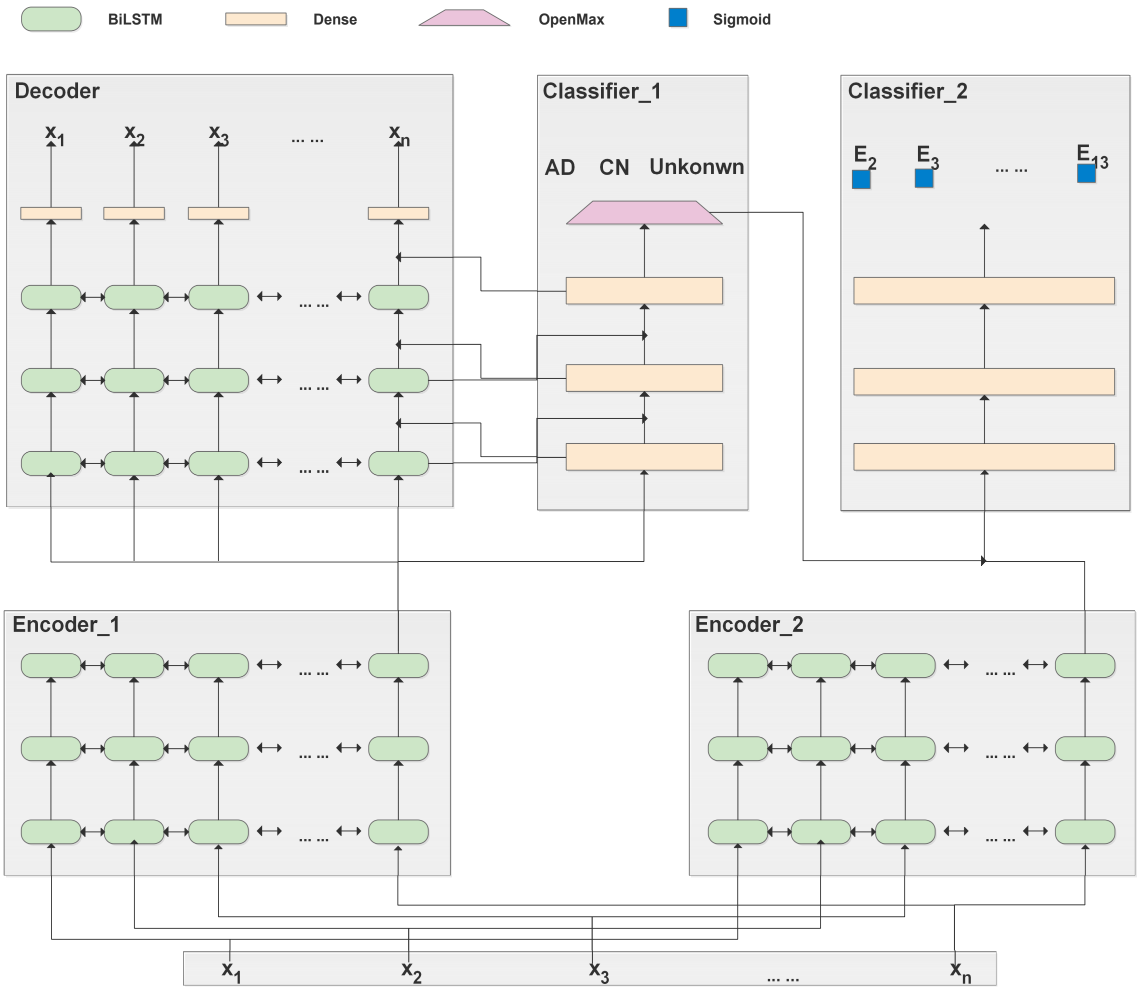


Fig. S 1: **The open, dynamic machine learning framework of OpenClinicalAI.** The OpenClinicalAI framework contains four independent modules and one accessory module. $Encoder\_1$ processes the input data for the $Classifier\_1$, and $Encoder\_2$ processes the input data for the $Classifier\_2$. The $Classifier\_1$ introduces the OpenMax mechanism to identify unknown categories of subjects. The accessory module $Decoder$ is used to help the $Classifier\_1$ retain features of the sample and improve the ability to identify unknown categories of subjects. The $Classifier\_2$ is used to select the examination to be carried out in the next step. In addition, the length of input data of the OpenClinicalAI framework is variable to adapt to data of different subjects at different visits.

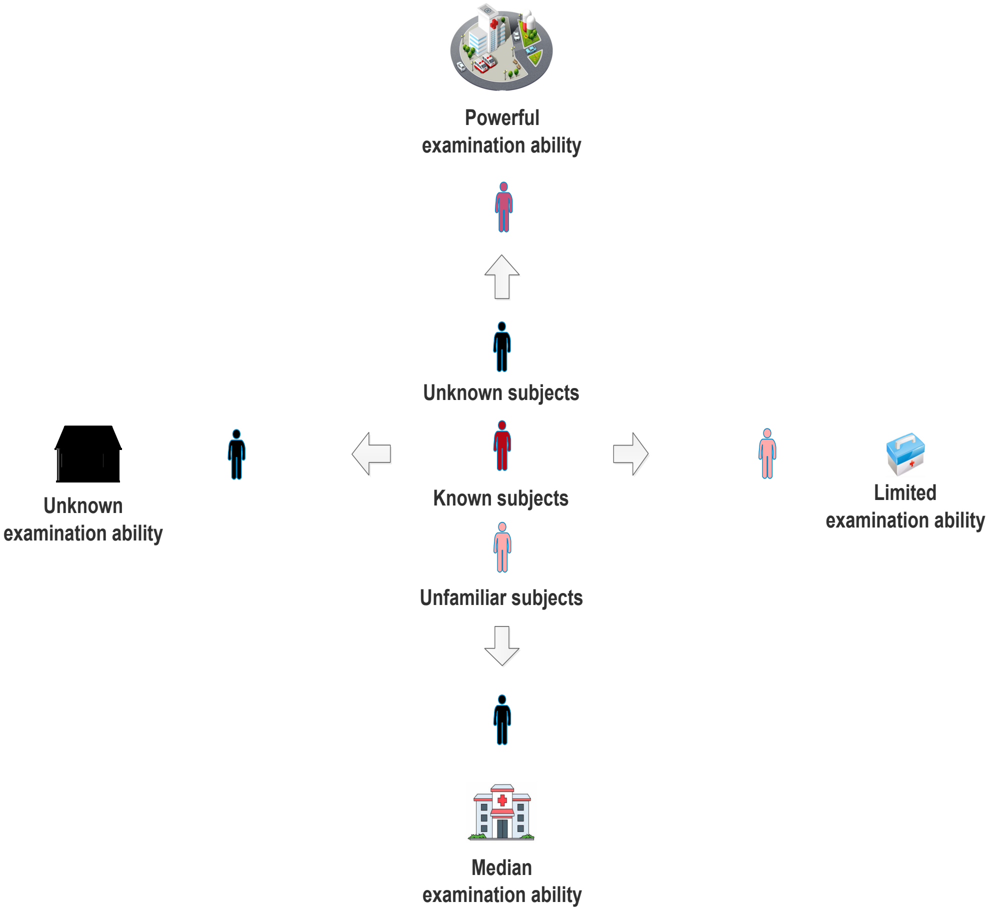


Fig. S 2: **The real-world setting of Clinical AIBench.** Subjects in real-world settings are different with various situations. They contain different pre-known categories and unknown and unfamiliar categories for the specific clinician or AI diagnostic system. The visit of subjects to a particular medical institution can not be pre-specified and hence are uncertain. Medical institutions in real-world settings also are different with different executive abilities of the examination. The executive ability of the examination in various medical institutions is very different from small-scale country clinics to large-scale hospitals. In addition, it is difficult to know by advance all the specific medical institutions that will deploy the AI system and their particular executive abilities of the examination.

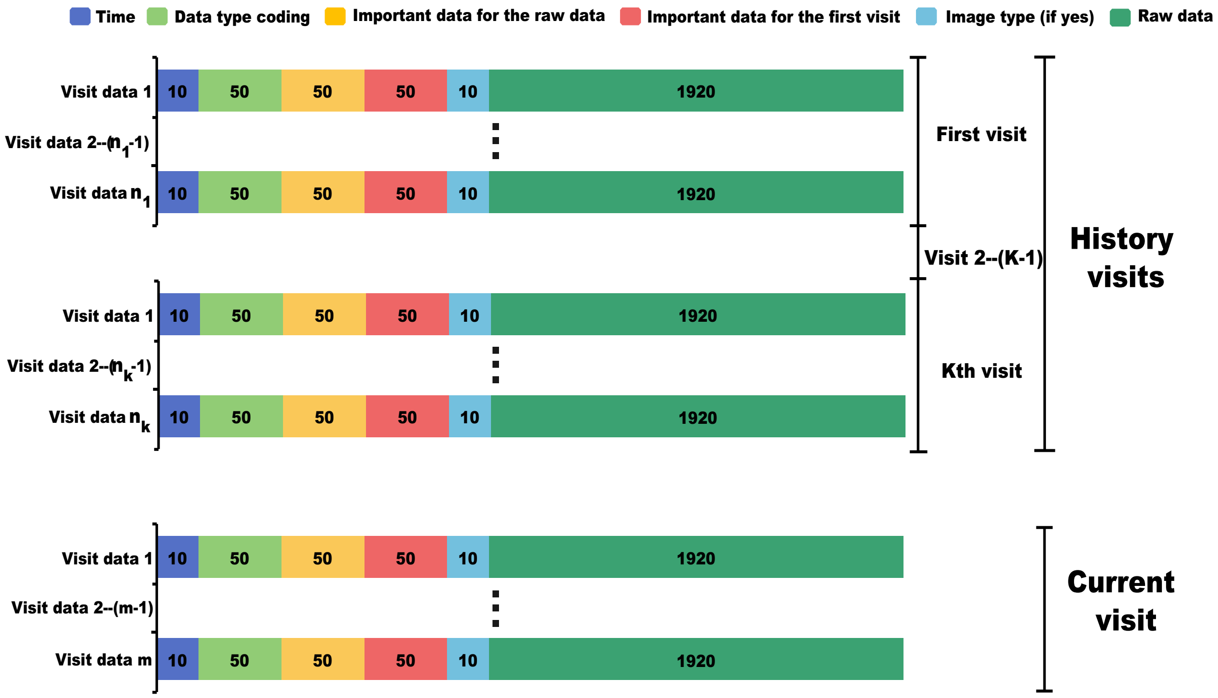


Fig. S 3: **Data framework for single subject.** Our data representation framework comprehensively considers the historical visit information and current visit information of the subject. The data with the earlier time is farther away from the current data.

Table S 1: **Characteristics of subjects.**

| | | Data set | Training set | Validation set | Test set |
|---|---|---|---|---|---|
| Age | 54-59.9 | 80 | 36 | 2 | 59 |
| | 60-69.9 | 596 | 246 | 10 | 442 |
| | 70-70.9 | 1048 | 528 | 46 | 695 |
| | 80-80.9 | 395 | 213 | 14 | 259 |
| | 90-91.9 | 6 | 1 | 1 | 4 |
| Gender | Female | 1130 | 560 | 44 | 785 |
| | Male | 997 | 465 | 29 | 675 |
| Educate | 4-7 | 11 | 4 | 0 | 8 |
| | 8-10 | 40 | 18 | 2 | 23 |
| | 11-13 | 353 | 176 | 13 | 243 |
| | 14-16 | 823 | 403 | 26 | 558 |
| | 17-20 | 900 | 424 | 32 | 628 |
| Ethnic category | Hisp/Latino | 73 | 32 | 5 | 49 |
| | Not Hisp/Latino | 2042 | 986 | 67 | 1404 |
| | Unknown | 12 | 7 | 1 | 7 |
| Racial category | Asian | 40 | 20 | 0 | 25 |
| | Black | 88 | 41 | 5 | 57 |
| | Hawaiian/Other PI | 2 | 0 | 0 | 2 |
| | More than one | 25 | 10 | 0 | 18 |
| | White | 1964 | 954 | 68 | 1350 |
| | Am Indian/Alaskan | 4 | 0 | 0 | 4 |
| | Unknown | 4 | 0 | 0 | 4 |
| Marriage | Married | 1618 | 805 | 59 | 1100 |
| | Never_married | 73 | 30 | 3 | 48 |
| | Widowed | 238 | 114 | 8 | 165 |
| | Divorced | 191 | 75 | 3 | 141 |
| | Unknown | 7 | 1 | 0 | 6 |
| Category | AD | 740 | 587 | 44 | 109 |
| | CN | 589 | 466 | 31 | 92 |
| | MCI | 1082 | 0 | 0 | 1082 |
| | SMC | 280 | 0 | 0 | 280 |

Table S 2: **The visit distribution of subjects.**

| Visit | Data set | Training set | Validation set | Test set |
|---|---|---|---|---|
| first visit | 2126 | 705 | 53 | 1368 |
| m06 | 1515 | 604 | 45 | 866 |
| m12 | 1475 | 621 | 43 | 811 |
| m18 | 329 | 79 | 5 | 245 |
| m24 | 1217 | 591 | 32 | 594 |
| m36 | 804 | 338 | 23 | 443 |
| m48 | 638 | 311 | 14 | 313 |
| m60 | 399 | 178 | 13 | 208 |
| m72 | 395 | 207 | 14 | 174 |
| m84 | 268 | 128 | 6 | 134 |
| m96 | 146 | 74 | 3 | 69 |
| m108 | 100 | 55 | 1 | 44 |
| m120 | 75 | 39 | 1 | 35 |
| m132 | 55 | 33 | 0 | 22 |
| m144 | 39 | 19 | 1 | 19 |
| m156 | 12 | 4 | 0 | 8 |

Table S 3: **The diagnosis strategies for the test set.**

| | Diagnosis strategies | | | | | | | | | | | | | Visit number of subject | | | |
|---|---|---|---|---|---|---|---|---|---|---|---|---|---|---|---|---|---|
| | Base | Cog | CE | Neur | FB | PE | Blood | Urine | MRI | FDG | AV45 | Gene | CSF | AD | CN | Unknown | Total |
| **1** | 1 | 1 | 0 | 0 | 0 | 0 | 0 | 0 | 0 | 0 | 0 | 0 | 0 | 244 | 280 | 2680 | 3204 |
| **2** | 1 | 1 | 1 | 1 | 1 | 1 | 0 | 0 | 1 | 0 | 0 | 0 | 0 | 16 | 23 | 697 | 736 |
| **3** | 1 | 1 | 1 | 1 | 1 | 1 | 0 | 0 | 1 | 0 | 1 | 0 | 0 | 3 | 10 | 81 | 94 |
| **4** | 1 | 1 | 1 | 1 | 1 | 1 | 0 | 0 | 1 | 1 | 1 | 0 | 0 | 6 | 8 | 124 | 138 |
| **5** | 1 | 1 | 1 | 0 | 0 | 0 | 0 | 0 | 0 | 0 | 0 | 0 | 0 | 8 | 47 | 43 | 98 |
| **6** | 1 | 1 | 1 | 1 | 1 | 1 | 0 | 0 | 0 | 0 | 0 | 0 | 0 | 10 | 6 | 183 | 199 |
| **7** | 1 | 1 | 1 | 1 | 0 | 0 | 0 | 0 | 0 | 0 | 0 | 0 | 0 | 1 | 2 | 27 | 30 |
| **8** | 1 | 1 | 1 | 1 | 1 | 1 | 0 | 0 | 0 | 1 | 0 | 0 | 0 | 1 | 2 | 11 | 14 |
| **9** | 1 | 1 | 1 | 1 | 1 | 1 | 0 | 0 | 0 | 0 | 1 | 0 | 0 | 1 | 1 | 16 | 18 |
| **10** | 1 | 1 | 1 | 1 | 1 | 1 | 0 | 0 | 1 | 1 | 0 | 0 | 0 | 2 | 6 | 232 | 240 |
| **11** | 1 | 1 | 1 | 1 | 1 | 1 | 0 | 0 | 0 | 1 | 1 | 0 | 0 | 0 | 3 | 16 | 19 |
| **12** | 1 | 1 | 1 | 1 | 1 | 0 | 0 | 0 | 0 | 1 | 1 | 0 | 0 | 0 | 0 | 1 | 1 |
| **13** | 1 | 1 | 1 | 1 | 1 | 0 | 0 | 0 | 0 | 0 | 0 | 0 | 0 | 1 | 0 | 33 | 34 |
| **14** | 1 | 1 | 0 | 1 | 1 | 1 | 0 | 0 | 1 | 1 | 1 | 0 | 0 | 0 | 0 | 2 | 2 |
| **15** | 1 | 1 | 0 | 1 | 1 | 1 | 0 | 0 | 1 | 0 | 0 | 0 | 0 | 0 | 0 | 1 | 1 |
| **16** | 1 | 1 | 1 | 1 | 1 | 1 | 1 | 0 | 1 | 1 | 1 | 1 | 1 | 2 | 0 | 113 | 115 |
| **17** | 1 | 1 | 1 | 1 | 1 | 1 | 1 | 1 | 0 | 0 | 0 | 0 | 0 | 5 | 14 | 10 | 29 |
| **18** | 1 | 1 | 1 | 1 | 1 | 1 | 0 | 0 | 1 | 0 | 0 | 1 | 0 | 0 | 3 | 21 | 24 |
| **19** | 1 | 1 | 1 | 1 | 1 | 1 | 1 | 0 | 1 | 1 | 1 | 1 | 0 | 3 | 0 | 13 | 16 |
| **20** | 1 | 1 | 1 | 1 | 1 | 1 | 1 | 0 | 0 | 0 | 0 | 0 | 0 | 1 | 0 | 42 | 43 |
| **21** | 1 | 1 | 1 | 1 | 1 | 1 | 1 | 0 | 1 | 0 | 0 | 0 | 0 | 0 | 1 | 2 | 3 |
| **22** | 1 | 1 | 1 | 1 | 1 | 1 | 0 | 0 | 1 | 0 | 1 | 1 | 0 | 0 | 5 | 21 | 26 |
| **23** | 1 | 1 | 1 | 1 | 1 | 1 | 0 | 0 | 1 | 1 | 0 | 1 | 0 | 1 | 0 | 9 | 10 |
| **24** | 1 | 1 | 1 | 1 | 1 | 1 | 1 | 1 | 1 | 1 | 0 | 1 | 1 | 0 | 0 | 54 | 54 |
| **25** | 1 | 1 | 1 | 1 | 1 | 1 | 1 | 0 | 1 | 0 | 0 | 1 | 0 | 0 | 0 | 60 | 60 |
| **26** | 1 | 1 | 1 | 1 | 1 | 1 | 1 | 1 | 1 | 0 | 0 | 1 | 1 | 0 | 0 | 67 | 67 |
| **27** | 1 | 1 | 1 | 1 | 1 | 1 | 1 | 0 | 1 | 1 | 0 | 1 | 0 | 0 | 0 | 55 | 55 |
| **28** | 1 | 1 | 1 | 1 | 1 | 1 | 1 | 1 | 1 | 1 | 0 | 1 | 0 | 0 | 0 | 1 | 1 |
| **29** | 1 | 1 | 1 | 1 | 1 | 1 | 1 | 1 | 1 | 0 | 0 | 1 | 0 | 0 | 0 | 3 | 3 |
| **30** | 1 | 1 | 1 | 1 | 1 | 1 | 1 | 0 | 1 | 1 | 0 | 1 | 1 | 0 | 0 | 2 | 2 |
| **31** | 1 | 1 | 1 | 1 | 1 | 1 | 0 | 0 | 1 | 1 | 1 | 1 | 0 | 0 | 0 | 13 | 13 |
| **32** | 1 | 1 | 1 | 1 | 1 | 1 | 1 | 0 | 1 | 1 | 1 | 0 | 0 | 0 | 0 | 1 | 1 |
| **33** | 1 | 1 | 1 | 1 | 1 | 1 | 1 | 0 | 1 | 0 | 0 | 1 | 1 | 0 | 0 | 1 | 1 |
| **34** | 1 | 1 | 1 | 1 | 1 | 1 | 0 | 0 | 1 | 0 | 0 | 1 | 1 | 0 | 0 | 1 | 1 |
| **35** | 1 | 1 | 1 | 1 | 1 | 1 | 0 | 0 | 0 | 0 | 1 | 1 | 0 | 0 | 0 | 1 | 1 |

Table S 4: **Medical institutions with different examination abilities in the test set.**

| | Medical institution without examination capabilities[1] | | | | | | | | | | | | | Visit number of subject in the condition of medical institution | | | |
|---|---|---|---|---|---|---|---|---|---|---|---|---|---|---|---|---|---|
| | Base | Cog | CE | Neur | FB | PE | Blood | Urine | MRI | FDG | AV45 | Gene | CSF | AD | CN | Unknown | Total |
| **1** | 0 | 0 | 0 | 0 | 0 | 0 | 1 | 1 | 0 | 0 | 0 | 0 | 0 | 23 | 35 | 952 | 1010 |
| **2** | 0 | 0 | 0 | 0 | 0 | 0 | 1 | 1 | 0 | 1 | 0 | 1 | 1 | 0 | 2 | 11 | 13 |
| **3** | 0 | 0 | 0 | 0 | 0 | 0 | 1 | 1 | 0 | 1 | 0 | 0 | 0 | 3 | 8 | 63 | 74 |
| **4** | 0 | 0 | 0 | 0 | 0 | 0 | 1 | 1 | 1 | 0 | 0 | 0 | 0 | 1 | 4 | 23 | 28 |
| **5** | 0 | 0 | 0 | 0 | 0 | 0 | 1 | 1 | 1 | 1 | 0 | 1 | 1 | 1 | 0 | 1 | 2 |
| **6** | 0 | 0 | 0 | 0 | 0 | 0 | 1 | 1 | 0 | 1 | 1 | 1 | 1 | 0 | 2 | 39 | 41 |
| **7** | 0 | 0 | 0 | 0 | 0 | 0 | 1 | 1 | 1 | 0 | 0 | 1 | 1 | 0 | 1 | 4 | 5 |
| **8** | 0 | 0 | 0 | 0 | 0 | 0 | 0 | 0 | 0 | 0 | 1 | 0 | 0 | 3 | 0 | 19 | 22 |
| **9** | 0 | 0 | 0 | 0 | 0 | 0 | 0 | 0 | 1 | 0 | 0 | 0 | 0 | 1 | 0 | 3 | 4 |
| **10** | 0 | 0 | 0 | 0 | 0 | 0 | 1 | 1 | 1 | 1 | 1 | 1 | 1 | 1 | 1 | 11 | 13 |
| **11** | 0 | 0 | 0 | 0 | 0 | 0 | 1 | 1 | 1 | 1 | 0 | 0 | 0 | 0 | 1 | 13 | 14 |
| **12** | 0 | 0 | 0 | 0 | 0 | 0 | 1 | 1 | 0 | 0 | 1 | 1 | 1 | 0 | 0 | 26 | 26 |
| **13** | 0 | 0 | 0 | 0 | 0 | 1 | 1 | 1 | 1 | 0 | 0 | 0 | 0 | 0 | 0 | 1 | 1 |
| **14** | 0 | 0 | 0 | 0 | 0 | 0 | 0 | 0 | 0 | 1 | 1 | 0 | 0 | 0 | 0 | 69 | 69 |
| **15** | 0 | 0 | 0 | 0 | 0 | 0 | 0 | 0 | 1 | 1 | 0 | 0 | 0 | 0 | 0 | 1 | 1 |
| **16** | 0 | 0 | 0 | 0 | 0 | 0 | 1 | 1 | 0 | 0 | 0 | 1 | 1 | 0 | 0 | 19 | 19 |
| **17** | 0 | 0 | 0 | 0 | 0 | 0 | 1 | 1 | 0 | 0 | 1 | 0 | 0 | 0 | 1 | 6 | 7 |
| **18** | 0 | 0 | 1 | 0 | 0 | 0 | 1 | 1 | 0 | 0 | 0 | 1 | 1 | 0 | 0 | 1 | 1 |
| **19** | 0 | 0 | 1 | 0 | 0 | 0 | 1 | 1 | 0 | 0 | 0 | 0 | 0 | 0 | 0 | 2 | 2 |
| **20** | 0 | 0 | 0 | 0 | 0 | 0 | 1 | 1 | 0 | 0 | 1 | 1 | 0 | 0 | 0 | 5 | 5 |
| **21** | 0 | 0 | 0 | 0 | 0 | 0 | 0 | 1 | 0 | 0 | 0 | 0 | 0 | 12 | 45 | 85 | 142 |
| **22** | 0 | 0 | 0 | 0 | 0 | 0 | 1 | 1 | 0 | 1 | 1 | 0 | 1 | 0 | 3 | 21 | 24 |
| **23** | 0 | 0 | 0 | 0 | 0 | 0 | 0 | 0 | 0 | 0 | 0 | 1 | 0 | 1 | 0 | 0 | 1 |
| **24** | 0 | 0 | 0 | 0 | 0 | 0 | 1 | 1 | 0 | 1 | 0 | 0 | 1 | 0 | 4 | 10 | 14 |
| **25** | 0 | 0 | 0 | 0 | 0 | 0 | 1 | 1 | 0 | 0 | 1 | 0 | 1 | 1 | 0 | 9 | 10 |
| **26** | 0 | 0 | 0 | 0 | 0 | 0 | 0 | 1 | 0 | 1 | 1 | 0 | 1 | 0 | 0 | 60 | 60 |
| **27** | 0 | 0 | 0 | 0 | 0 | 0 | 0 | 1 | 0 | 0 | 1 | 0 | 1 | 0 | 0 | 54 | 54 |
| **28** | 0 | 0 | 0 | 0 | 0 | 0 | 0 | 0 | 0 | 0 | 1 | 0 | 1 | 0 | 0 | 1 | 1 |
| **29** | 0 | 0 | 0 | 0 | 0 | 0 | 0 | 0 | 0 | 1 | 1 | 0 | 1 | 0 | 0 | 3 | 3 |
| **30** | 0 | 0 | 0 | 0 | 0 | 0 | 0 | 1 | 0 | 0 | 0 | 0 | 1 | 0 | 0 | 5 | 5 |
| **31** | 0 | 0 | 0 | 0 | 0 | 0 | 1 | 1 | 0 | 0 | 0 | 0 | 1 | 0 | 0 | 9 | 9 |
| **32** | 0 | 0 | 0 | 0 | 0 | 0 | 0 | 1 | 0 | 1 | 1 | 0 | 0 | 0 | 0 | 1 | 1 |
| **33** | 0 | 0 | 0 | 0 | 0 | 0 | 1 | 1 | 0 | 1 | 1 | 0 | 0 | 0 | 0 | 1 | 1 |
| **34** | 0 | 0 | 0 | 0 | 0 | 0 | 1 | 1 | 0 | 1 | 1 | 1 | 0 | 0 | 0 | 2 | 2 |
| **35** | 0 | 0 | 0 | 0 | 0 | 0 | 0 | 0 | 0 | 0 | 1 | 1 | 0 | 0 | 0 | 5 | 5 |
| **36** | 0 | 0 | 0 | 0 | 0 | 0 | 0 | 0 | 0 | 1 | 0 | 0 | 0 | 0 | 0 | 1 | 1 |
| **37** | 0 | 0 | 0 | 0 | 0 | 0 | 0 | 1 | 0 | 0 | 1 | 0 | 0 | 0 | 0 | 1 | 1 |
| **38** | 0 | 0 | 0 | 0 | 0 | 0 | 1 | 0 | 0 | 1 | 0 | 0 | 0 | 0 | 0 | 1 | 1 |
| **39** | 0 | 0 | 0 | 0 | 0 | 0 | 1 | 1 | 1 | 1 | 0 | 0 | 1 | 0 | 0 | 1 | 1 |
| **40** | 0 | 0 | 0 | 0 | 0 | 0 | 1 | 0 | 0 | 1 | 1 | 0 | 0 | 0 | 0 | 1 | 1 |

[1] The examination is marked as 1, meaning that the medical institution cannot perform this examination for the subject. The examination is marked as 0, indicating that (1) the medical institution can perform this examination for the subject, or (2) OpenClinicalAI does not request for performing this examination during the diagnosis of the subject though the medical institution may not be able to perform this examination for the subject. It is worth noting that the examination ability in the test set may be different from other AI systems since 0 may mean that OpenClinicalAI does not request for performing this examination during the diagnosis of the subject. However, the medical institution may not be able to perform this examination for the subject.

Table S 5: **SNPs relate to AD.**

| SNP_NAME | SNP_NAME | SNP_NAME | SNP_NAME |
|---|---|---|---|
| rs429358 | rs7412 | rs10948363 | rs7274581 |
| rs17125944 | rs4147929 | rs6656401 | rs11771145 |
| rs6733839 | rs983392 | rs10498633 | rs28834970 |
| rs9271192 | rs35349669 | rs9331896 | rs1476679 |
| rs10792832 | rs2718058 | rs190982 | rs10838725 |
| rs11218343 | rs4844610 | rs10933431 | rs9271058 |
| rs75932628 | rs9473117 | rs12539172 | rs10808026 |
| rs73223431 | rs3740688 | rs7933202 | rs3851179 |
| rs17125924 | rs12881735 | rs3752246 | rs6024870 |
| rs7920721 | rs138190086 | rs4723711 | rs4266886 |
| rs61822977 | rs6733839 | rs10202748 | rs115124923 |
| rs115675626 | rs1109581 | rs17265593 | rs2597283 |
| rs1476679 | rs78571833 | rs12679874 | rs2741342 |
| rs7831810 | rs1532277 | rs9331888 | rs7920721 |
| rs3740688 | rs7116190 | rs526904 | rs543293 |
| rs11218343 | rs6572869 | rs12590273 | rs7145100 |
| rs74615166 | rs2526378 | rs117481827 | rs7408475 |
| rs3752246 | rs7274581 | | |

Table S 6: **The normal range of indicators.**

| | | AD_Normal | | CN_Normal | |
|---|---|---|---|---|---|
| | | Low | High | Low | High |
| Medical history | Psychiatric | 0 | 0 | 0 | 0 |
| | Neurologic (other than AD) | 0 | 0 | 0 | 0 |
| Symptoms[1] | Present_count_21[2] | 0 | 6 | 0 | 6 |
| | Present_count_28[3] | 0 | 8 | 0 | 8 |
| Cognitive Change Index[4] | Score_12[5] | 32.2188 | 60 | 12 | 13.5634 |
| | Score_20[6] | 50.3438 | 100 | 20 | 22.0845 |
| CDRSB[7] | | 2 | 18 | 0 | 0 |
| Alzheimer's Disease Assessment Scale[8] | ADAS11[9] | 10 | 70 | 0 | 11.264 |
| | ADAS13[10] | 18 | 85 | 0 | 17.67 |
| | ADASQ4 | 5 | 10 | 0 | 6 |
| MMSE[11] | | 0 | 27 | 25 | 30 |
| MOCA[12] | | 0 | 23 | 26 | 30 |
| Preclinical Alzheimer's Cognitive Composite[13] | mPACCdigit | -30.0745 | -7.6955 | -5.1733 | 4.7304 |
| | mPACCtrailsB | -29.7277 | -6.7798 | -4.8523 | 4.3338 |

[1] Nausea, Vomiting, Diarrhea, Constipation, Abdominal discomfort, Sweating, Dizziness, Low energy, Drowsiness, Blurred vision, Headache, Dry mouth, Shortness of breath, Coughing, Palpitations, Chest pain, Urinary discomfort (e.g., burning), Urinary frequency, Ankle swelling, Muscloskeletal pain, Rash, Insomnia, Depressed mood, Crying, Elevated mood, Wandering, Fall, Other.

[2] Nausea to Rash

[3] Nausea to Other

[4] The CCI scale is in `https://adni.bitbucket.io/reference/cci.html`.

[5] CCI1 to CCI12

[6] CCI1 to CCI20

[7] The CDR scale is in `https://adni.bitbucket.io/reference/cdr.html`.

[8] The Alzheimer's Disease Assessment Scale-Cognitive scale is in `https://adni.bitbucket.io/reference/adas.html`.

[9] Q1 to Q11

[10] Q1 to Q13

[11] The Mini Mental State Exam scale is in `https://adni.bitbucket.io/reference/mmse.html`.

[12] The Montreal Cognitive Assessment scale is in `https://adni.bitbucket.io/reference/moca.html`.

[13] The calculation method of Preclinical Alzheimer's Cognitive Composite is in `https://ida.loni.usc.edu/pages/access/studyData.jsp?categoryId=16&subCategoryId=43`.

**Algorithm S 1 The examination label algorithm.**

**Input:** The label set $y_{true}$, the prediction set $y_{pred}$, diagnosis strategy set $exam\_strategy$ for a subject in a visit.

**Output:** Next examination set $next\_exam$

Sort the $exam\_strategy$ by the number of examinations in a diagnosis strategy.
**for** $i = 0 \quad to \quad len$(exam_strategy) **do**
  **for** $j = i + 1 \quad to \quad len$(exam_strategy) **do**
    **if** $exam\_strategy$[i]$\subset$$exam_strategy$[j] **then**
      $gain = sum(y_{true}[j] \times y_{pred}[j] - y_{true}[i] \times y_{pred}[i]) + sum(\sim y_{true}[i] \times y_{pred}[i] - \sim y_{true}[j] \times y_{pred}[j])$
      **if** $gain > 0$ **then**
        The next examination of current examination strategy $exam\_strategy$[i] is label by $exam\_strategy$[j].
      **end if**
    **end if**
  **end for**
**end for**

**Algorithm S 2 The modified OpenMax algorithm.**

**Input:** The abnormal pattern dataset $X$, the FitHigh function from libMR (*63*), the MiniBatchKMeans function from scikit-learn (*70*), the number of the center of known categories of subject $N$, quantiles $Q$.

**Output:** The centers of known categories of subject $C$, and libMR models $Model$, the threshold of known categories of subject $Thr$.

$X[i]$ is the abnormal pattern dataset of $ith$ known categories of subject, in which every data $x \in X$ is belong to $ith$ known categories of subject and is correctly classified by the trained AI model. $L$ is the number of the known categories of subject.
**for** $i = 0 \quad to \quad (L-1)$ **do**
    $C[i] = MiniBatchKMeans(X[i], N[i])$
**end for**
$Dist = []$
**for** $i = 0 \quad to \quad (L-1)$ **do**
    **for** $x \quad in \quad X[i]$ **do**
        $Dist[i]$.add(distance(x,$C[i]$,$C_{others}$)    // $distance = sqrt(min\_distance(x, C[i])^2 + (1 - min\_distance(x, C_{others}))^2)$
    **end for**
**end for**
**for** $i = 0 \quad to \quad (L-1)$ **do**
    $Model[i]$=FitHigh($Dist[i]$)
    $Thr[i]$ is the $Q[i]$ quantile of the $Dist[i]$
**end for**
Return $C$, $Model$, $Thr$

**Algorithm S 3 OpenMax probability estimation.**

**Input:** Abnormal pattern of the subject $X = \{x_1, x_2, ..., x_n\}$, raw data of subject $Z$, activation vector $V(Z) = \{v_1(Z), v_2(Z)\}$, The centers of known categories of subject $C$, and libMR models $Model$, the threshold of known categories of subject $Thr$, flag $F$, the numer of top classes to revise $\alpha$.

**Output:** The prediction probability $\hat{P}$.

$L$ is the number of the known categories of subject.
Let $s(i) = argsort(v_j(Z))$
Let $Dist = []$
**for** $i = 0 \quad to \quad (L-1)$ **do**
  $dist[i] = distance(X, C[i], C_{others})$
**end for**
**for** $i = 1 \quad to \quad \alpha$ **do**
  $\omega_i(Z) = 1 - \frac{\alpha - i}{\alpha} * models[i-1].w\_score(\text{dist}[i-1])$
**end for**
Revise activation vector $\hat{V}(Z) = V(Z) \circ \omega(Z)$
Define $\hat{v}_0(Z) = \sum_i v_i(Z)(1 - \omega_i(Z))$
$\hat{P}(y = j \mid Z) = \frac{e^{\hat{v}_j(Z)}}{\sum_{i=0}^{2} e^{\hat{v}_i(Z)}}$
**if** $F$ **then**
  $abnor\_score = []$
  **for** $j = 1 \quad to \quad (L-1)$ **do**
    $diff = dist[j-1] - thr[j-1]$
    **if** $diff <= 0$ **then**
      $abnor\_score$.append(0)
    **else**
      $tmp\_abnor\_score = diff/thr[j-1]$
      **if** $tmp\_abnor\_score > 1$ **then**
        $tmp\_abnor\_score = 1$
      **end if**
      $abnor\_score$.append($tmp\_abnor\_score$)
    **end if**
  **end for**
  **for** $j = 1 \quad to \quad (L-1)$ **do**
    $\hat{P}(y = j \mid Z) = \hat{P}(y = j \mid Z) * (1 - abnor\_score[j-1])$
  **end for**
  $\hat{P}(y = 0 \mid Z) = 1 - \sum_{j=1}^{L-1} \hat{P}(y = j \mid Z)$
**end if**
Return $\hat{P}$

**Algorithm S 4 The prediction algorithm.**

**Input:** The base information $data_{base}$ and history recodes $data_h$ for a subject in a visit, the trained model $model$. The threshold $\delta$, and $\gamma$.

**Output:** The label of the subject.

$data_{input}$=$data_h$ concatenates $data_{base}$<br>
**while** True **do**<br>
    $result_{pred}$, $next\_examination_{pred}$=$model.predict(data_{input})$<br>
    **for** $i = 0 \quad to \quad len(result_{pred})$ **do**<br>
        **if** $result_{pred}[i] >= \delta[i]$ **then**<br>
            Return i // When $i == len(result_{pred}) - 1$, the result is representing unknown<br>
        **end if**<br>
    **end for**<br>
    $is\_concat\_new\_data = False$<br>
    **for** $i = 0 \quad to \quad len(next\_examination_{pred})$ **do**<br>
        **if** $next\_examination_{pred}[i] >= \gamma[i]$ **then**<br>
            **if** The $ith$ examination is able to execute by medical institution **then**<br>
                $data_{input}$=$data_{input}$ concat $data_{ith}$<br>
                $is\_concat\_new\_data = True$<br>
            **end if**<br>
        **end if**<br>
    **end for**<br>
    **if** not $is\_concat\_new\_data$ **then**<br>
        Select a less cost and common examination $jth$ examination which do not execute in this visit and is able to execute by medical institution.<br>
        **if** $jth$ examination is selected **then**<br>
            $data_{input}$=$data_{input}$ concat $data_{jth}$<br>
            $is\_concat\_new\_data = True$<br>
        **end if**<br>
    **end if**<br>
    **if** not $is\_concat\_new\_data$ **then**<br>
        Return unknown<br>
    **end if**<br>
**end while**

### References

48. R. C. Petersen, *et al.*, *Neurology* **74**, 201 (2010).

49. M. W. Weiner, *et al.*, *Alzheimer's & Dementia* **6**, 202 (2010).

50. M. W. Weiner, *et al.*, *Alzheimer's & Dementia* **11**, 865 (2015).

51. M. W. Weiner, *et al.*, *Alzheimer's & Dementia* **13**, 561 (2017).

52. S. Darkner, Fdg-pet template mni152 1mm (2013).

53. S. M. Smith, *et al.*, *Neuroimage* **23**, S208 (2004).

54. N. Seneca, C. Burger, I. Florea pp. 1–3 (2011).

34. G. Huang, Z. Liu, L. Van Der Maaten, K. Q. Weinberger, *Proceedings of the IEEE conference on computer vision and pattern recognition* (2017), pp. 4700–4708.

55. J.-C. Lambert, *et al.*, *Nature genetics* **45**, 1452 (2013).

56. B. W. Kunkle, *et al.*, *Nature genetics* **51**, 414 (2019).

57. R. S. Desikan, *et al.*, *PLoS medicine* **14**, e1002258 (2017).

16. C. Geng, S.-j. Huang, S. Chen, *IEEE transactions on pattern analysis and machine intelligence* (2020).

58. P. Perera, *et al.*, *Proceedings of the IEEE/CVF Conference on Computer Vision and Pattern Recognition* (2020), pp. 11814–11823.

11. A. Bendale, T. Boult, *Proceedings of the IEEE conference on computer vision and pattern recognition* (2015), pp. 1893–1902.

59. A. Kendall, Y. Gal, R. Cipolla, *Proceedings of the IEEE conference on computer vision and pattern recognition* (2018), pp. 7482–7491.

14. H. Lee, *et al.*, *Nature biomedical engineering* **3**, 173 (2019).

60. Y. Zhang, Q. V. Liao, R. K. Bellamy, *Proceedings of the 2020 Conference on Fairness, Accountability, and Transparency* (2020), pp. 295–305.

61. P. Linardatos, V. Papastefanopoulos, S. Kotsiantis, *Entropy* **23**, 18 (2021).

62. Z. Ge, S. Demyanov, Z. Chen, R. Garnavi, *British Machine Vision Conference 2017* (British Machine Vision Association and Society for Pattern Recognition, 2017).

63. W. J. Scheirer, A. Rocha, R. Michaels, T. E. Boult, *IEEE Transactions on Pattern Analysis and Machine Intelligence (PAMI)* **33**, 1689 (2011).

64. C. R. Jack Jr, *et al.*, *Alzheimer's & dementia* **7**, 257 (2011).

65. R. A. Sperling, *et al.*, *Alzheimer's & dementia* **7**, 280 (2011).

66. M. S. Albert, *et al.*, *Alzheimer's & dementia* **7**, 270 (2011).

67. M. C. Donohue, *et al.*, *JAMA neurology* **71**, 961 (2014).

68. D. P. Kingma, J. Ba, *ICLR (Poster)* (2015).

69. B. Efron, R. J. Tibshirani, *An introduction to the bootstrap* (CRC press, 1994).

70. D. Sculley, *Proceedings of the 19th international conference on World wide web* (2010), pp. 1177–1178.
